# High-Power Short-Duration vs Low-Power Long-Duration Ablation for Pulmonary Vein Isolation: A Substudy of the AWARE Randomized Controlled Trial

**DOI:** 10.1101/2023.06.16.23291526

**Authors:** Jacqueline Joza, Girish M Nair, David H Birnie, Pablo B Nery, Calum J Redpath, Jean-Francois Sarrazin, Jean Champagne, Jean-Francois Roux, Charles Dussault, Ratika Parkash, Martin Bernier, Laurence D. Sterns, John Sapp, Paul Novak, George Veenhuyzen, Carlos A. Morillo, Sheldon M. Singh, Mouhannad M. Sadek, Mehrdad Golian, Andres Klein, Marcio Sturmer, Vijay S. Chauhan, Paul Angaran, Martin S. Green, Jordan Bernick, George A Wells, Vidal Essebag, the AWARE - RCT (Augmented Wide Area Circumferential Catheter Ablation for Reduction of Atrial Fibrillation Recurrence)

## Abstract

**Background:** Pulmonary vein isolations (PVI) are being performed using a high-power, short duration (HPSD) strategy. The purpose of this study was to compare the clinical efficacy and safety outcomes of a HPSD vs low-power long duration (LPLD) approach to PVI in patients with paroxysmal atrial fibrillation (AF).

**Methods:** Patients were grouped according to a HPSD (≥40 W) or LPLD (≤ 35 W) strategy. The primary endpoint was the one-year recurrence of any atrial arrhythmia lasting ≥ 30 seconds, detected using three 14-day ambulatory continuous ECG monitoring. Procedural and safety endpoints were also evaluated. The primary analysis were regression models incorporating propensity scores yielding adjusted relative risk (RR_a_) and mean difference (MD_a_) estimates.

**Results:** Of the 398 patients included in the AWARE Trial, 173 (43%) underwent HPSD and 225 (57%) LPLD ablation. The distribution of power was 50 W in 75%, 45 W in 20% and 40 W in 5% in the HPSD group, and 35W with 25W on the posterior wall in the LPLD group. The primary outcome was not statistically significant at 30.1% vs 22.2% in HPSD and LPLD group with RR_a_ 0.77 (95% confidence interval [CI]) 0.55-1.10; p=0.165). The secondary outcome of repeat catheter ablation was not statistically significant at 6.9% and 9.8% (RR_a_ 1.59 [95% CI 0.77-3.30]; p=0.208) respectively. The incidence of any ECG documented AF during the blanking period was numerically lower in the HPSD group: 1.7% vs 8.0% (RR_a_ 3.95 [95% CI 1.00-15.61; p=0.049). The total procedure time was significantly shorter in the HPSD group (MD_a_ 97.5 minutes [95% CI 84.8-110.4)]; p<0.0001) with no difference in adjudicated serious adverse events.

**Conclusions:** A HPSD strategy was associated with significantly shorter procedural times with similar efficacy in terms of clinical arrhythmia recurrence. Importantly, there was no signal for increased harm with a HPSD strategy.

**Graphical Abstract:** 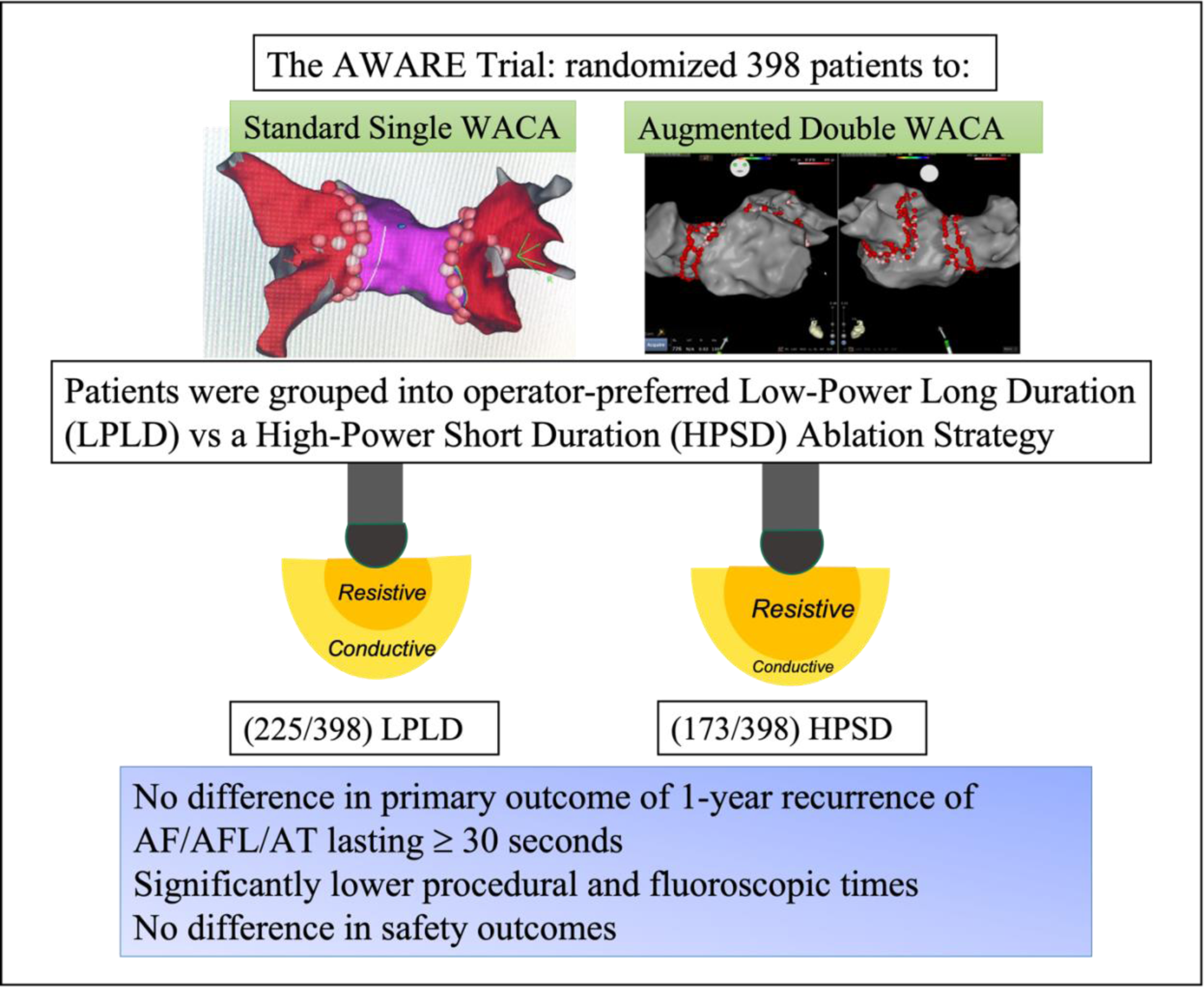

**Non-standard Abbreviations and Acronyms:** HPSD: High-Power Short Duration; LPLD: Low-Power Long Duration; QOL: Quality of Life; WACA: wide area circumferential ablation; PVI: Pulmonary Vein Isolation; AF: Atrial Fibrillation

**Clinical Perspective:** *What is known:* -The optimal power and duration of ablation lesions to produce durable pulmonary vein isolation remain unclear. -Nonrandomized studies have suggested clinical efficacy with high-power short duration radiofrequency ablation vs low-power long duration.

*What this study adds:* -In this large substudy of the AWARE Trial, a high-power short duration radiofrequency ablation strategy was found to be similarly effective as a low-power long duration strategy with no difference in time to first recurrence of any AF lasting ≥ 30 seconds. -Procedural were substantially reduced with high-power short duration ablation with no significant difference in overall complication rates.

## Introduction

Novel ablation techniques for pulmonary vein isolation (PVI) have been of significant interest, but no more so than that of the high-power, short-duration (HPSD) lesion strategy which has seen an incredible uptake among experienced and new operators alike. Its theoretical advantages of a shorter procedure time, increased likelihood of first-pass isolation, theoretically improved safety profile, and ease of applicability have made it an attractive alternative to a standard low-power, long-duration lesion (LPLD) application.

The biophysics profile of HPSD support its use in the atria. In vitro modeling has confirmed a linear relationship between increasing total energy delivery and lesion depth, where the energy delivered is dependent both on power and time (Energy (J) = Power (W) x time (seconds)) ^1, 2^. However, lesion *width* appears to be larger with HPSD ablation for the same energy delivered because it creates a large zone of direct resistive heating with shorter temperature decay time ^3^. This has been confirmed in a porcine animal model demonstrating improved lesion-to-lesion uniformity with wider lesions at similar depths as compared to LPLD lesions ^4^. This has translated into higher acute success rates with shorter radiofrequency (RF) time and lower incidence of chronic pulmonary vein (PV) reconnections in observational studies ^5^.

A comparable safety profile to conventional ablation has also been observed across observational studies ^5–7^. Parameters identifying durable lesion formation with simultaneous lesion safety have been adapted to HPSD. These include predefined impedance drops, loss of local electrograms, unipolar signal modification, and proprietary algorithms such as lesion size index (LSI) and ablation index (AI) ^3^ both of which include power in its calculation (as opposed to the Force Time Integral [FTI] which does not). HPSD does not appear to increase the risk of atrio-esophageal fistula where appropriate titration of contact force and lesion duration were instituted on the posterior wall ^8–10^. On the contrary, a large observational study suggested a lower rate of atrio-esophageal fistula with HPSD compared to use of 35W on the posterior wall ^9^.

The Standard Versus Augmented Ablation of Paroxysmal Atrial Fibrillation for Reduction of Atrial Fibrillation Recurrence (AWARE) Study (ClinicalTrials.gov Identifier: NCT02150902) randomized 398 patients to receive RF catheter ablation for PVI with either a standard single wide area circumferential ablation (WACA) or an augmented doubled WACA^11^. In this multicenter Canadian study, approximately half of the Canadian AWARE investigators routinely used a HPSD strategy for PVI. This substudy compares ablation outcomes in patients receiving HPSD as compared to LPLD ablation in the context of a randomized controlled trial.

## Methods

The AWARE Trial design and methodology has been previously described ^12^. Briefly, 398 symptomatic and drug refractory patients with paroxysmal atrial fibrillation (AF) were randomized to receive radiofrequency catheter ablation for PVI with either a standard single wide area circumferential ablation (WACA) or an augmented double WACA. The hypothesis was that a secondary barrier and wider area of atrial ablation would increase the chance of durable PVI, thereby resulting in reduction of atrial arrhythmias, without affecting procedural safety. The primary outcome was atrial arrhythmia (atrial tachycardia, atrial flutter, or atrial fibrillation ≥ 30 seconds) recurrence between 91- and 365-days post ablation, off antiarrhythmic drugs. Secondary outcomes included need for repeat catheter ablation, procedural and safety variables. Monitoring was performed through use of three 14-day ambulatory continuous ECG monitoring at 3-, 6-, and 12-months post ablation, and three clinical visits. All AF-related hospital visits and any ECG documented atrial arrhythmias during patient encounters were also used for determining the primary outcome.

All patients were randomized after the first WACA lesion set was completed and prior to evaluation of PV isolation. If randomized to the single WACA arm, PV isolation with entrance and exit block was confirmed. After an observation period of 20 minutes, adenosine was given, and subsequent ablation performed as needed. In patients randomized to the double WACA arm, a second lesion set was delivered, followed by a 20-minute observation time after the initial WACA, and then testing for PV bidirectional block. The Biosense Webster Inc. SMARTTOUCH TC or SMARTTOUCH SF ablation catheter were used for ablation. Midway through the trial, the protocol was modified to allow the option of high-power short duration (HPSD) RF delivery.

This AWARE substudy examined trial outcomes based on an operator preferred HPSD or LPLD PVI strategy across 10 centers in Canada. The substudy population included all randomized AWARE patients. HPSD was defined as a power setting of ≥ 40 W, and LPLD was defined as a power of ≤ 35W. For both HPSD and LPLD effective ablation lesion tags were represented on the electroanatomic map using the VISITAG (Biosense Webster Inc) software with ablation lesions >2mm apart with an interlesion distance (ILD) of 5mm. In the LPLD group, an FTI ≥ of 400 g/second (g/s) or an AI of ≥ 550 was recommended on the anterior, inferior, and superior aspects of the PV antra. A maximum power of 25 watts was permitted along the posterior wall of the LA, with a recommended FTI of 300 g/s but not > 400g/s, or a target AI of ≥ 400. Operators using the HPSD approach were permitted to use RF energy from 40 to 50 W and energy delivery duration ranging from 5 to 15 seconds during ablation. It was recommended to target a minimum contact force (CF) of 5 g and not to exceed a CF of 20 g during ablation. Ablation lesions delivered on the posterior wall were delivered for shorter duration and/or lower CF and a lower CF and/or duration advised if the catheter tip was close to the esophagus or phrenic nerve. Given the lack of validation of FTI or AI using *high power* settings, lesions did not have prespecified FTI or AI targets.

Data was analyzed according to HPSD or LPLD ablation strategy. Categorical variables were presented as frequencies and percentages, and continuous variables as the mean and standard deviation (SD), as well as median and inter-quartile range (IQR). The two strategies were compared using the standardized difference, with the absolute value of <0.10 indicating a clinically minimal difference^13^. Since the ablation strategies are subgroups of the AWARE Trial, and not determined by randomization an adjusted analysis was needed. The propensity score incorporating all the baseline patient characteristics was computed and the inverse probability treatment weight (IPTW) method using propensity scores was used for all analyses. For the primary outcome of the 1-year recurrence rate of any ECG documented AF, AFl or AT between the two strategies, the Poisson regression model with robust error variances was used and the adjusted relative risk (RR_a_) estimate and the corresponding 95% confidence interval (CI) were calculated^14^ or the secondary categorical outcomes, a similar analysis plan was followed. For the continuous outcomes, such as procedure duration and fluoroscopy duration, a multiple linear regression model was used for comparing the two strategies, and the adjusted least squares mean difference (MD_a_) estimate and the corresponding 95% CI were calculated. The primary outcome was compared between key subgroups chosen based on clinical relevance (i.e., sex, age, duration of AF, congestive heart failure (CHF), hypertension, diabetes, stroke history, CHA_2_DS_2_VAS_c_, WACA (AWARE trial randomization arms single vs double) and based on the test of the ablation strategy by subgroup interaction term in the logistic regression model. The ablation strategies will be compared for all primary and secondary outcomes within the single and double WACA therapy arms. Statistical analysis was performed using SAS software version 9.4 (SAS Institute). The two-sided nominal p-value < 0.05 was considered statistically significant.

The AWARE Trial funding was provided by the Canadian Institutes of Health Research, the University of Ottawa Heart Institute (UOHI) and Biosense Webster Inc., Canada.

## RESULTS

### Patient Population

Of the 398 patients included in the AWARE Trial, 173 (43%) underwent a HPSD ablation strategy and 225 (57%) a LPLD ablation strategy. The HPSD group consisted of patients with a mean age of 62 years and 40% female, whereas the LPLD group was younger 60 (SD 0.23) with only 28% female (SD 0.26) (Table 1). Other differences included: more hypertension (47% vs 37%, SD 0.21) and diabetes mellitus (12% vs 8%, SD 0.12) in the HPSD group, a higher CHA_2_DS_2_-VASc score in the HPSD group (1.75 v 1.30, SD 0.35) and less use of baseline anticoagulants medication (78% vs 86%, SD 0.20) and aspirin/antiplatelet agents (6% VS 16%, SD 0.33) and more ARB (20% VS 7%, SD 0.38) in the HPSD group. The HPSD and LPLD groups were similar on other characteristics including: BMI CIED, time since diagnosis, prior history of stroke/TIAs or thromboembolism, antiarrhythmic drugs (43% in each group), left atrial diameter and LAVI (all SD<0.10). After adjusting for the propensity scores, all absolute standardized differences are < 0.10 indicating similar baseline characteristics when the IPTW are used (Table 1).

**Table 1.** Baseline characteristics: Comparison of the low-power long duration (LPLD) and high-power short duration (HPSD) ablation strategies

|  | LPLD Group<br>N=225 | HPSD Group<br>N=173 | Absolute Standardized<br>Difference |  |
| --- | --- | --- | --- | --- |
|  |  |  | Unadjusted | Adjusted* |
| Age (years) | 60.10± 9.46 | 62.20± 8.99 | 0.228 | 0.021 |
| Sex | 62(27.6%) | 69(39.9%) | 0.263 | 0.010 |
| BMI (Kg/m<sup>2</sup>) | 28.86±5.33 | 29.36±6.01 | 0.088 | 0.020 |
| CIED | 2(0.89%) | 3(1.73%) | 0.074 | 0.017 |
| Time since<br>diagnosis (months) | 54.69± 59.41 | 50.93± 65.59 | 0.060 | 0.015 |
| CHF | 6(2.7%) | 5(2.9%) | 0.013 | 0.021 |
| Hypertension | 83(36.9%) | 82(47.4%) | 0.214 | 0.021 |
| Diabetes Mellitus | 19(8.4%) | 21(12.2%) | 0.122 | 0.008 |
| Prior History of<br>Stroke/TIA or<br>thromboembolism | 14(6.2%) | 13(7.5%) | 0.051 | 0.010 |
| CHADS<sub>2</sub>-65 Score |  |  |  |  |
| CHA<sub>2</sub>DS<sub>2</sub>-VASc<br>score | 1.30± 1.25 | 1.75± 1.35 | 0.346 | 0.011 |
| Antiarrhythmic |  |  |  |  |
| Propafenone | 16(7.11%) | 4(2.31%) | 0.228 | 0.085 |
| Flecainide | 56(24.9%) | 44(25.4%) | 0.012 | 0.020 |
| Sotalol | 22(9.8%) | 18(10.4%) | 0.021 | 0.014 |
| Amiodarone | 32(14.2%) | 29(16.8%) | 0.070 | < 0.001 |
| Dronedarone | 3(1.33%) | 3(1.73%) | 0.033 | 0.002 |
| No antiarrhythmic<br>medications | 96(42.7%) | 75(43.4%) | 0.014 | 0.044 |
| ACE-Inhibitors | 35(15.6%) | 27(15.6%) | 0.001 | 0.006 |
| ARB | 16(7.1%) | 19(19.9%) | 0.383 | 0.004 |
| Beta-blockers | 106(47.1%) | 82(47.4%) | 0.006 | 0.033 |
| Diltiazem/Verapamil | 35(15.6%) | 29(16.7%) | 0.033 | 0.009 |
| Digoxin | 2(0.89%) | 4(2.31%) | 0.113 | 0.031 |
| Aspirin/antiplatelet<br>agents | 36(16.0%) | 10(5.8%) | 0.333 | 0.097 |
| Statins | 64(28.4%) | 71(41.0%) | 0.267 | 0.025 |
| Anticoagulants |  |  |  |  |
| No systemic<br>anticoagulation prior<br>to enrolment | 32(14.2%) | 38(21.0%) | 0.202 | 0.055 |
| Warfarin | 5(2.22%) | 2(1.16%) | 0.082 | 0.017 |
| Dabigatran | 18(8.0%) | 4(2.31%) | 0.259 | 0.026 |
| Rivaroxaban | 91(40.44%) | 54(31.21%) | 0.193 | 0.002 |
| Apixaban | 76(33.78%) | 65(37.57%) | 0.079 | 0.036 |
| Edoxaban | 3(1.33%) | 10(5.78%) | 0.242 | 0.026 |
| Left atrial diameter (mm) | 41.93± 35.03<br>N=147 | 41.78± 33.51<br>N=98 | 0.013 | 0.020 |
| LAVI (ml/m<sup>2</sup>) | 32.88± 10.17<br>N=160 | 33.12± 10.06<br>N=115 | 0.024 | 0.004 |

\*Adjusted using propensity scores with inverse probability treatment weighting
**Legend:** Plus-minus values are provided as mean ± SD and other data presented as number (percentage). AAD = antiarrhythmic drug; ACE= angiotensin-converting enzyme; AF=atrial fibrillation; ARB=Angiotensin receptor blocker; BMI= body mass index (weight in kilograms divided by square of the height in meters); CHF=congestive heart failure (New York Heart Association Heart Failure class II or Left ventricular ejection fraction < 50%); CIED= cardiac implantable electronic device; LA=left atrium; LAVI= Left atrial volume index; TIA=transient ischemic attack.

### Procedural characteristics

In the HPSD group, the distribution of power used on the anterior wall was 50 W in 75%, 45 W in 20%, and 40 W in 5%. The distribution on the posterior wall was 50 W in 70%, 45 watts in 25%, 40 watts in 3% (5 patients), 35 watts in 0.6% (1 patient), and 25 watts in 1.2% (2 patients). The distribution on the roof was 50 watts in 73%, 45 watts in 23%, 40 watts in 2.6% (4 patients), and 25 watts in 0.6% (1 patient). The median lesion duration was 10 sec (IQR 10,10) anteriorly, 8 sec (IQR 7,8) on the posterior wall, and 9 seconds (IQR 8,10) on the roof with an average contact force of 10, 10, and 9 respectively on each location. The power delivery in the LPLD group was limited to 35 W on the anterior, superior, and inferior aspects of the LA antrum, and to a maximum of 25 W at all posterior antral sites. Data on average contact force in the LPLD group was not collected.

### Clinical Outcomes

The primary outcome of the 1-year recurrence rate of any ECG documented AF, AFL or AT (symptomatic or asymptomatic lasting ≥ 30 seconds) after a blanking period of 90 days occurred in 52 (30.1%) in patients in the HPSD group and 50 (22.2%) in the LPLD group and was not statistically significant (RR_a_ 0.77 [95% CI, 0.55-1.11]; p=0.165) (Table 2). The secondary arrhythmia outcome of need for repeat catheter ablation because of documented recurrence of *symptomatic* arrhythmia trended in favor of the HPSD occurring in 12 (6.9%) of patients compared to 22 (9.8%) in the LPLD group but was not statistically significant (RR_a_ 1.59 [95% CI, 0.77-3.30]; p=0.208) in the HPSD and LPLD group respectively. Any ECG documented AF (symptomatic or asymptomatic) during the blanking period (within 90 days post procedure) was significantly lower in the HPSD group occurring in 3 (1.7%) patients versus 18 (8%) in the LPLD group (RR_a_ 3.95 [95% CI, 1.00-5.61]; p=0.049). The presence of an atrial arrhythmia in the blanking period predicting the primary outcome was 67% (3 blanking period arrhythmias predicting 2 primary outcomes) in the HPSD group, and 44% (18 blanking period arrhythmias predicting 8 primary outcomes) in the LPLD group.

**Table 2.**
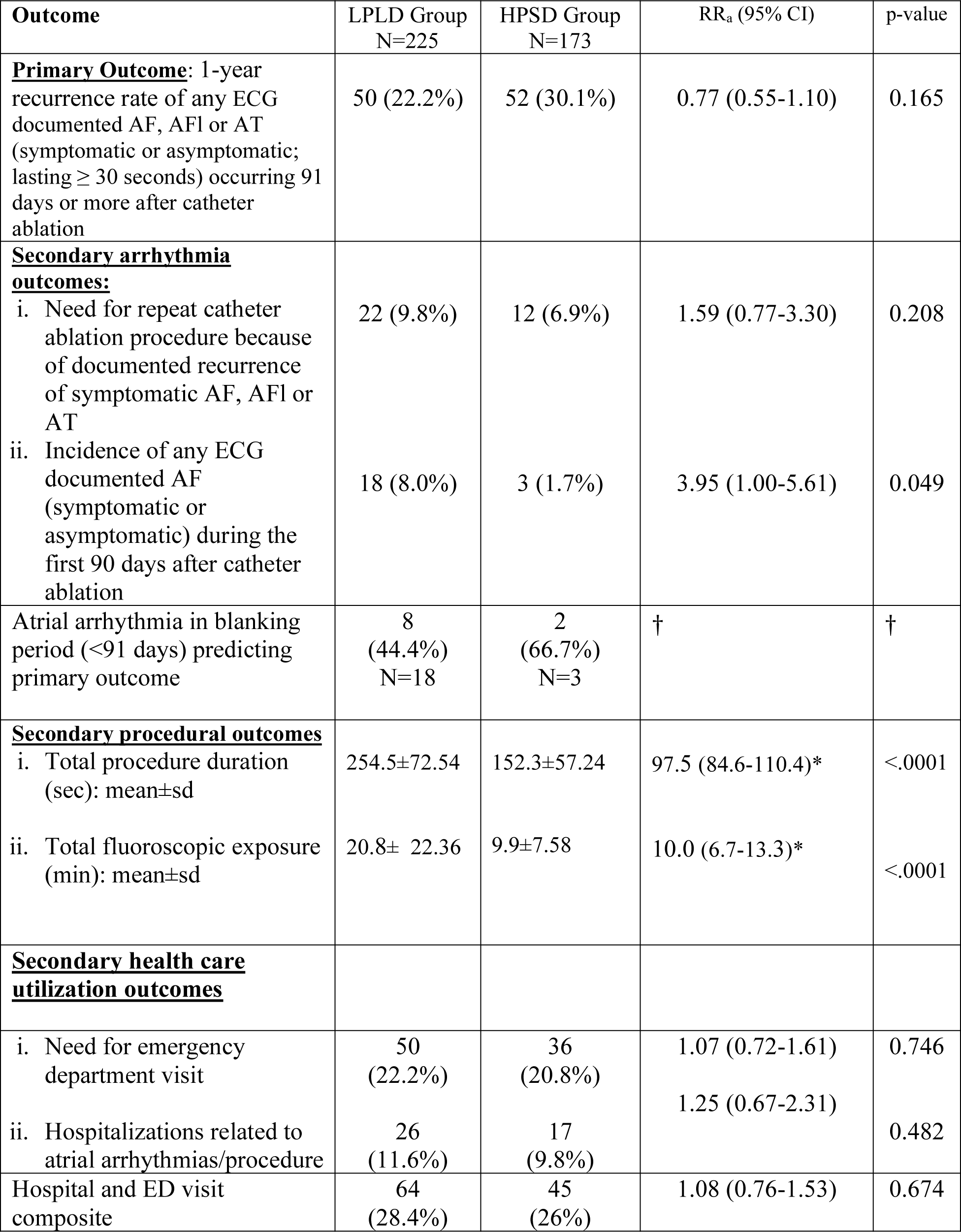
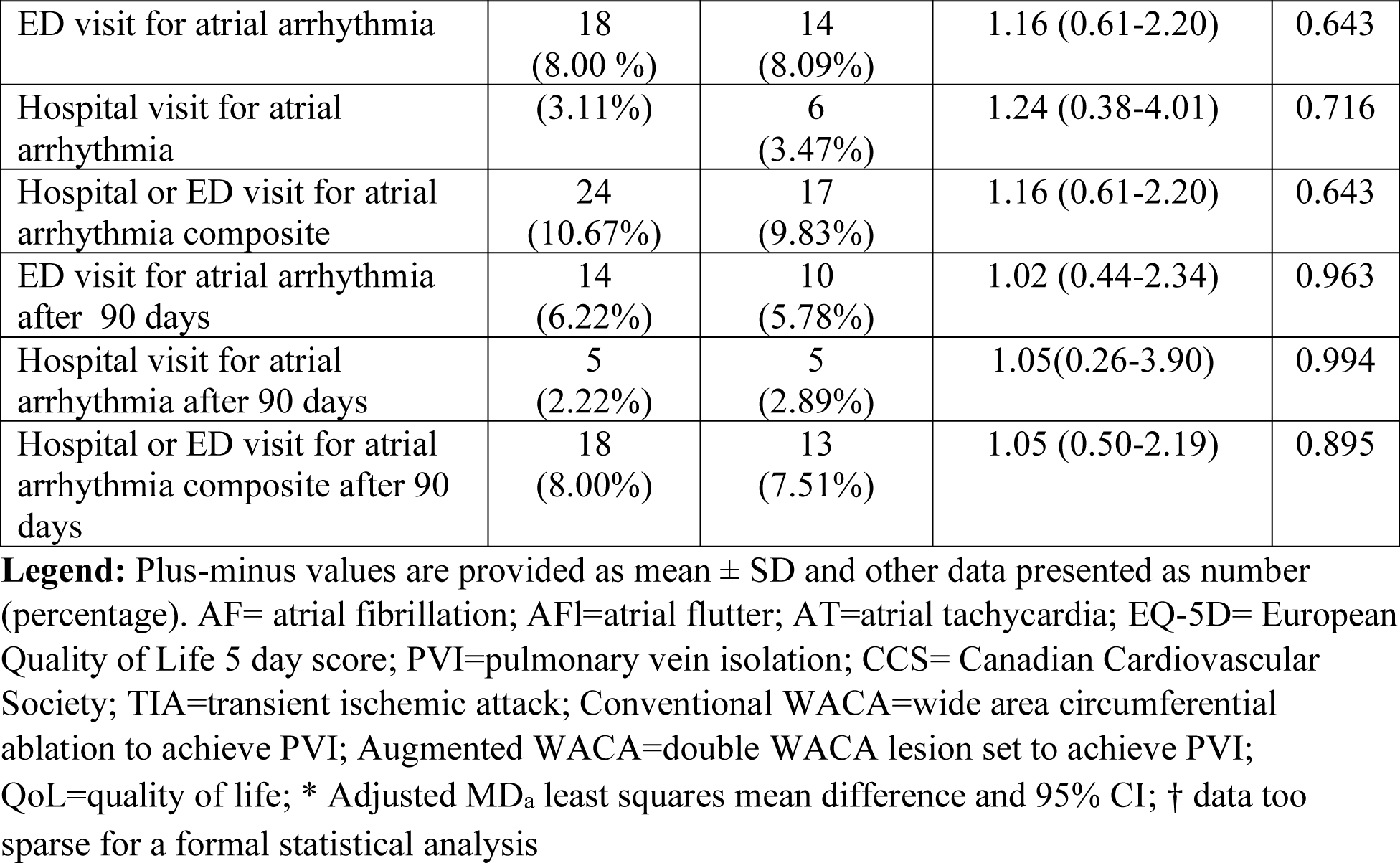
Primary and secondary clinical and procedural outcomes: Comparison of the low-power long duration (LPLD) and high-power short duration (HPSD) ablation strategies

There were no significant differences in secondary health care utilization outcomes or need for emergency department visits or hospitalizations related to a recurrent arrhythmia or from a complication of the procedure itself during the follow-up and blanking periods (Table 2). In a sensitivity analysis examining the use of 45 W vs 50 W in the HPSD group, the primary outcome was not significantly different with 22.9% (8/35) events occurring in the 45 W group vs 33% (42/128) in the 50 W group (RR_a_ 0.69; 95% CI, 0.36-1.34, p=0.305).

The ablation strategies were compared on the primary outcome across subgroups of sex, age (≤ 65, >65 years), duration of AF (<50, ≥ 50 months), CHF, hypertension, diabetes, stroke history and CHA_2_DS_2_VAS_c_ (<4, ≥ 4), and no significant differences were identified for the ablation strategy by subgroup interaction. (Table 3) The p-values for interaction were all >0.05, except for the duration of AF (p=0.0272) and hypertension (p=0.0291) for which the duration of AF <50 months and no hypertension subgroups had significant reduction in the 1-year recurrence rate.

**Table 3.**
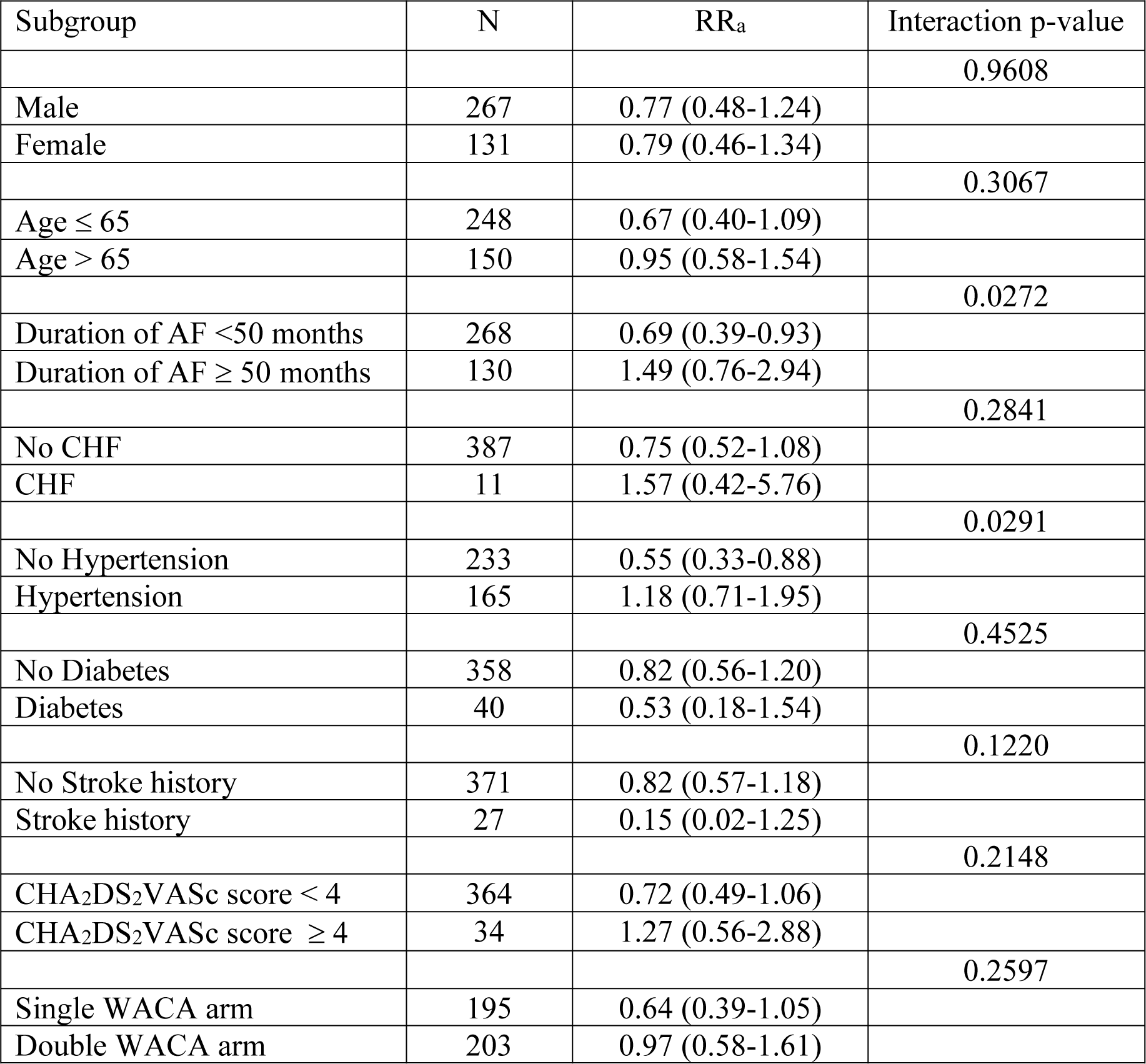
One-year recurrence rate: Comparison of the low-power long duration (LPLD) and high-power short duration (HPSD) ablation strategies by subgroups

| Subgroup | N | RR<sub>a</sub> | Interaction p-value |
| --- | --- | --- | --- |
|  |  |  | 0.9608 |
| Male | 267 | 0.77 (0.48-1.24) |  |
| Female | 131 | 0.79 (0.46-1.34) |  |
|  |  |  | 0.3067 |
| Age ≤ 65 | 248 | 0.67 (0.40-1.09) |  |
| Age > 65 | 150 | 0.95 (0.58-1.54) |  |
|  |  |  | 0.0272 |
| Duration of AF <50 months | 268 | 0.69 (0.39-0.93) |  |
| Duration of AF ≥ 50 months | 130 | 1.49 (0.76-2.94) |  |
|  |  |  | 0.2841 |
| No CHF | 387 | 0.75 (0.52-1.08) |  |
| CHF | 11 | 1.57 (0.42-5.76) |  |
|  |  |  | 0.0291 |
| No Hypertension | 233 | 0.55 (0.33-0.88) |  |
| Hypertension | 165 | 1.18 (0.71-1.95) |  |
|  |  |  | 0.4525 |
| No Diabetes | 358 | 0.82 (0.56-1.20) |  |
| Diabetes | 40 | 0.53 (0.18-1.54) |  |
|  |  |  | 0.1220 |
| No Stroke history | 371 | 0.82 (0.57-1.18) |  |
| Stroke history | 27 | 0.15 (0.02-1.25) |  |
|  |  |  | 0.2148 |
| CHA<sub>2</sub>DS<sub>2</sub>VASc score < 4 | 364 | 0.72 (0.49-1.06) |  |
| CHA<sub>2</sub>DS<sub>2</sub>VASc score ≥ 4 | 34 | 1.27 (0.56-2.88) |  |
|  |  |  | 0.2597 |
| Single WACA arm | 195 | 0.64 (0.39-1.05) |  |
| Double WACA arm | 203 | 0.97 (0.58-1.61) |  |

### Procedural Outcomes

The procedural outcomes were all significantly lower in the HPSD group (Table 2). In particular, the mean difference in the total procedure duration was significantly shorter (MD_a_ 97.5 [95% CI, 84.6 -110.4]; p<0.0001). It is important to note the presence of a mandatory 20-minute waiting time after the entrance and exit block were confirmed with adenosine (single WACA group, and after initial pass, double WACA group). The fluoroscopic exposure time was significantly higher in the LPLD group (MD_a_ 10.0 [95% CI, 6.7-13.3]; p<0.0001).

### Safety Outcomes

There was no significant difference in the rate of adjudicated serious adverse events between the HPSD and LPLD groups (Table 4). Procedural complications included cardiac perforation with tamponade in 2 (0.89%) patients in the LPLD group compared to 0 in the HPSD, major bleeding requiring transfusion in 1 (0.44%) patient with 2 (0.89%) minor bleeding events in the LPLD group compared to 0 in the HPSD group, congestive heart failure in 2 (0.89%) patients in the LPLD group vs 1 (0.58%) in the HPSD, and 1 (0.58%) patient with pulmonary vein stenosis in the HPSD group as compared to 0 in the LPLD group, which occurred in the double WACA group. One perforated esophageal ulcer was noted in the LPLD group which occurred in a chronically immunosuppressed patient on high-dose corticosteroids and a biologic immunosuppressant for nephrotic syndrome. One pseudoaneurysm was noted in each group, and 1 arterio-venous fistula in the LPLD group.

**Table 4.** Serious adverse events: Comparison of the low-power long duration (LPLD) and high-power short duration (HPSD) ablation strategies

| Serious Adverse Events | LPLD Group<br>N=225 | HPSD Group<br>N=173 |
| --- | --- | --- |
|  | n (%) | n (%) |
| Death | 0 | 0 |
| Atrial Fibrillation | 3 (1.33%) | 0 |
| Atrial Flutter | 1 (0.44%) | 1 (0.58%) |
| Congestive Heart Failure | 2 (0.89%) | 1 (0.58%) |
| Sinus bradycardia | 0 | 1(0.58%) |
| Ischemic Stroke | 0 | 0 |
| Transient ischemic attack | 0 | 0 |
| Major Bleeding (requiring transfusion) | 1 (0.44%) | 0 |
| Minor Bleeding | 2 (0.89 %) | 0 |
| Hypotension | 1 (0.44%) | 0 |
| Pneumonia | 1 (0.44%) | 0 |
| Pseudonaneurysm | 1 (0.44%) | 1 (0.58%) |
| Arterio-venous Fistula | 1 (0.44%) | 0 |
| Cardiac Perforation (with tamponade) | 2 (0.89%) | 0 |
| Pericarditis | 0 | 1 (0.58%) |
| Pulmonary Vein Stenosis | 0 | 1 (0.58%) |
| Diaphragmatic paralysis | 0 | 0 |
| Esophageal ulcer perforation | 1 (0.44%) | 0 |

A serious adverse event was defined as an adverse event that led to death; that led to a serious deterioration in health resulting in a life-threatening illness or injury, permanent impairment of a body structure or a body function, inpatient hospitalization, prolonged hospitalization (>24 hours), or medical or surgical experiment to prevent life-threatening illness, injury, or permanent impairment to a body structure or a body function; or that led to fetal distress, fetal death, or a congenital abnormality or birth defect.

### Outcomes of HPSD vs LPLD based on randomization group (single vs double WACA)

#### Single WACA arm

Of the 195 patients included in the single WACA arm of the AWARE trial, 83 (43%) underwent a HPSD ablation strategy and 112 (57%) a LPLD ablation strategy. The primary outcome in the single WACA group was not statistically significant occurring in 28 (39%) of patients in the HPSD group compared to 24 (21%) in the LPLD group (RR_a_ 0.64 [95% CI 0.39-1.05]; p=0.079) (Table S1). There were numerically fewer patients needing a repeat catheter ablation procedure with 5 (6%) and 14 (12%) patients in the HPSD and LPLD groups respectively (RR_a_ 2.66 [95% CI 0.94-7.53]; p=0.065). There were smaller numbers of documented asymptomatic or symptomatic AF episodes during the blanking period at 2 (2.4%) versus 12 (10.7%) patients in the HPSD and LPLD groups respectively but the adjusted comparison was not significant (RR_a_ 3.43 [95% CI 0.69-16.89]; p=0.130). Procedural and fluoroscopic times were significantly shorter in the HPSD arm, MD_a_ 96.8 ([95% CI 79.3-114.4]; p<0.0001) and MD_a_ 11.4 ([95% CI (6.65-16.14)]; p<0.0001) respectively.

#### Double WACA arm

Of the 203 patients included in the double WACA arm of the AWARE trial, 90 (44%) underwent a HPSD ablation strategy and 113 (56%) a LPLD ablation strategy. The primary outcome in the double WACA group was not statistically significant occurring in 24 (27%) and 26 (23%) of patients in the HPSD and LPLD groups respectively (RR_a_ 0.97 [95% CI 0.58-1.61]; p=0.902) (Table S2). The need for repeat catheter ablation was also not significantly different at 7 (7.8%) and 8 (7.1%) in the HPSD and LPLD groups respectively (RR_a_ 0.89 [95% CI 0.31-2.55]; p=0.834). The incidence of documented AF in the blanking period was numerically lower in the HPSD group but did not reach statistical significance 1 (1.1%) vs 6 (5.3%) patients) (RR_a_ 6.69 95% CI 0.79-55.79]; p=0.080). Procedural and fluoroscopic times were significantly shorter in the HPSD arm, MD_a_ 98.2 ([95% CI 79.0-117.3]; p<0.0001) and MD_a_ 8.5 ([95% CI 3.9-13.2]; p=0.0004) respectively.

## DISCUSSION

To our knowledge, this is the largest randomized trial to evaluate patients according to high power short duration vs low power long duration for AF ablation. This substudy reached several important conclusions: 1. A HPSD strategy was found to be similarly effective as a LPLD strategy with no difference in the clinical outcome using a highly sensitive definition of 1-year recurrence of *any* AF lasting ≥ 30 seconds. 2. There was no signal for increased harm with a HPSD strategy, and although there were no statistically significant differences in complication rates, there were numerically higher numbers of tamponades and the presence of a perforated esophageal ulcer in the LPLD group. 3. Procedural and fluoroscopy times were substantially reduced, approaching half the time for HPSD than for LPLD. This study was also hypothesis generating: 1. A trend towards a reduction in blanking period arrhythmias was noted in the HPSD group and appears to be consistent with other studies. Given that the absolute time required for catheter stability is shorter with HPSD ablation, this may have led to a higher proportion of RF lesions causing *irreversible* injury as compared to LPLD ablation accounting for this observation. It is also interesting that LPLD was associated with a higher rate of early recurrence of AF, and further research would be required to determine whether this might be related to a greater inflammatory response due to lower power ablation lesions. 2. There were numerically but not statistically significant lower numbers of repeat ablation procedures in the HPSD group. Presumably a repeat ablation would have been performed in the presence of symptom recurrence or new persistent AF. A lower number of repeat ablations in the HPSD group would therefore suggest improved clinically meaningful long-term durability as compared to LPLD. Longer follow-up would be necessary to determine a difference.

The overall success rates of PVI for paroxysmal AF range from 60-80% after a single procedure. Despite the advent of irrigated and contact force sensing catheters, PV reconnection occurs in 22% of PVs acutely and 15% of PVs 3 months after isolation ^15, 16^. Attempts to improve clinical outcomes have targeted the efficacy of lesion formation including a focus on catheter stability with steerable sheaths, atrial or ventricular pacing, and low-tidal volume ventilation settings/high-frequency jet ventilation ^17^. The adoption of real time lesion surrogates such as AI and LSI to demonstrate lesion quality have also advanced the field. The ‘CLOSE’-guided PVI protocol with an ILD of 6mm with a target AI of 400 posteriorly and 550 anteriorly was proposed and validated, demonstrating long-term durability with a single-procedure freedom from any atrial arrhythmia of 87% at 1 year and 78% at 2 years on no antiarrhythmic therapy ^18^. Additional electrogram (egm) monitoring beyond the ‘close’-guided PVI suggested to be of limited value based on characterization of real-time changes in bipolar and unipolar egms ^19^.

Based on the premise of wider lesion size with shorter energy delivery times, a HPSD strategy had the promise of even further improved outcomes. Irrigated catheters allowed for application of higher powers without the creation of steam pops; irrigation with 17ml/min allowed delivery of higher powers of 40 W up to 30 seconds as compared to 10 seconds with a lower irrigation flow at 2ml/min ^20^. Initial *in vitro* and *in vivo* ovine models compared standard settings of 40 W/30 sec with a temperature limit of 50 deg C with 40, 50, 60, 70, and 80 W for 5 second duration with saline irrigation set at 30ml/min ^2^. Compared with conventional settings, 50 and 60 W for 5 seconds achieved transmurality and was safer (steam pops were absent with HPSD ablations). Operators have investigated AI-guided PVI with different power protocols including 50 W anterior/40 W posterior, 40 W anterior/30 W posterior, and 30 W anterior/20 W posterior with the same target AI (400 anteriorly, 360 posteriorly and 260 on the esophagus) ^21^. Higher power applications significantly increased the first-pass isolation rate (85%, 80%, and 55% respectively), but no significant difference was noted in the AF-free survival rates at 6 months. Yavin *et al* compared 112 patients undergoing HPSD ablations to historical controls undergoing standard ablation ^5^. A higher acute success was seen with HPSD ablation (90.2% vs 83%, p=0.06) with shorter ablation times (17.2 +-3.4 minutes vs 31.1 +-5.6 minutes, p<0.001) and a lower incidence of chronic PV reconnections (16.6% vs 52.2%, p=0.03). Interestingly, in a higher proportion of HPSD applications, catheter motion was <1mm during 50% or more of the application duration, thereby improving stability during lesion delivery. Chen *et al* reported on their series of 122 consecutive patients undergoing HPSD ablation with 50 W targeting an AI of 550 anteriorly and 400 posteriorly with an ILD of 6mm ^22^. First-pass isolation was noted in 96.7%. Follow-up with 72-hour holters demonstrated a single-procedure freedom from clinical recurrence of atrial arrhythmia off antiarrhythmic drug of 85.2% (89% for paroxysmal AF and 80% for persistent AF) at 15 months. The mean contact force, RF duration, AI, and impedance drop at the anterior/posterior wall were 26 ± 14g/23 ± 12g, 16.2±7.5s/8.8 ± 3.6s, 552 ± 53/438± 47, and 13 ± 6Ω/9 ± 5Ω. Finally, a recent meta-analysis using a HPSD definition of > 40 W with ablation duration of 2-10 sec per site identified 6 prospective studies, 3 retrospective, and 1 randomized controlled trial. In 2467 patients, pooled analyses demonstrated higher rates of first-pass isolation than LPLD ablation (RR 1.2 [95% CI, 1.1-1.31]; P<0.001) and a reduction in atrial arrhythmia recurrence (RR 0.73 [95% CI, 0.58-0.91]; P=0.005) ^23^. Major complications and esophageal thermal injury were similar between groups.

Atrio-esophageal fistula as a consequence of LA ablation remains rare but devastating. Given that HPSD creates broader and shallower lesions with less conductive heating ^24^, it follows that a theoretical advantage may be a lower risk of atrio-esophageal fistula. In cadaver studies, the mean (min, max) LA transmural thickness at the anterior wall, roof, and posterior wall was found to be 1.86 ± 0.59 mm (0.6, 2.6), 1.06± 0.49 mm (0.8,1.51), and 1.4 ±. 0.46 mm (0.9, 2). Although a formalin effect appeared to influence the posterior wall measurements ^25^ these thicknesses correlated with measurements on computational tomography ^26^. The LA wall has also been reported to be thinner in patients with AF compared with those without ^27^. Other studies have revealed that the esophageal wall was <5mm from the endocardium in 40% of patients, and that the LA thickness differed significantly among several different regions of the posterior wall ^28^. Using same-day late gadolinium enhancement magnetic resonance imaging, moderate-to-severe esophageal enhancement was seen in 14.3% of patients undergoing AF ablation with both HPSD (50W/5s) and LPLD (<35W for 10-30 seconds).

HPSD has demonstrated clinical safety in our as well as in several other studies. Winkle *et al.* demonstrated posterior wall applications using 45-50 W for 2-10 seconds to be safe ^9^. In a study including 10,284 patients from four centers, one atrioesophageal fistula (0.0087%) occurred in 11,436 HPSD ablations performed using 45-50 W for 2-10 seconds on the posterior wall, while three atrioesophageal fistulas (0.12%) occurred in 2538 LPLD ablations using 35 W on the posterior wall for 20 seconds (p=0.021) ^10^. Notably, two of the three fistulas in the LPLD group did not undergo esophageal temperature monitoring. A strategy of HPSD with 45 W vs LPLD with 35 W using the ‘close’ protocol was evaluated in a prospective, randomized controlled study involving a total of 100 patients ^29^. Endoscopic evaluation performed in a proportion of patients revealed the presence of an ulcerative perforation in the HPSD group which required endoscopic stenting with normalization after 4 months. A superficial ulcerative lesion in a control group patient was treated conservatively, however both occurred following excessive AI applications (up to 460 and 480 respectively) with excessive contact force (average 30 g with peaks up to 50g). Finally, in another study of 122 patients undergoing HPSD ablation, patients with an esophageal temperature of ≥39 degrees C underwent post-ablation endoscopy. Three (2.5%) patients had asymptomatic endoscopic small erosion/erythema esophageal lesions with no serious adverse events observed ^30^.

This study represented a substudy of a randomized controlled trial with standardized protocols and uniform follow-up however the allocation of HPSD vs LPLD was not randomized and was according to operator preference. Data on the duration of the recurrent atrial arrhythmia or the presence or absence of symptoms during AF episodes during follow-up were not collected. This prevented understanding the indication to proceed with a repeat intervention or determine if the type of recurrence was clinically relevant. Data on first-pass isolation was not collected.

In this substudy of the AWARE trial, a HPSD ablation approach was found to be associated with a marked reduction in procedural duration and fluoroscopy time, while being similarly effective as a LPLD approach for the incidence of recurrent atrial arrhythmia post ablation for paroxysmal AF. A numerically lower number of blanking period arrhythmias were noted with HPSD with trends towards lower repeat ablations. Further studies with longer follow-up are necessary to confirm these findings.

## Data Availability

Data is available upon request

## Acknowledgements

none

## Sources of Funding

The AWARE RCT was funded by grants from the Canadian Institutes of Health Research, the University of Ottawa Heart Institute, Division of Cardiology and Academic Medical Organization, and by an unrestricted research grant from Biosense Webster (IIS-381). Dr. Essebag is supported by a Clinical Research Scholar Award from the Fonds de Recherche du Quebec Santé (FRQS).

## Disclosures

**Dr. Jacqueline Joza** reports investigator-initiated external grant support from Medtronic Inc. and consulting fees from Boston Scientific (Modest)and honoraria from Biosense Webster Canada (Modest). **Dr. Girish M. Nair** reports honoraria, speaking fees and grant support from Biosense Webster Inc. and Boston Scientific Inc. related to Atrial Fibrillation (Modest). **Dr. Pablo B. Nery** reports honoraria, speaking fees and grant support from Biosense Webstern Canada, not related to this work (Modest). **Dr. David H. Birnie** reports grants from Boehringer Ingelheim, Germany, grants from Pfizer and Bristol-Myers Squibb, New York (Modest). **Dr. George Veenhuyzen** has received honoraria & consulting fees from Medtronic, BMS-Pfizer, Servier, & Biotronik. **Dr. Jean-Francois Sarrazin** has received consulting fees from Biosense Webster. **Dr. Jean-Francois Roux** has received consulting feeds from Biosense Webster. **Dr. Carlos A. Morillo** has received honoraria/consulting fees from Abbott, Biosense Webster, Boston Scientific, and Medtronic for AF related lectures and research support. **Dr. Ratika Parkash** has received consulting fees/honoraria and research support from Abbott, Biosense Webster and Medtronic Inc. **Dr. John Sapp** has received honoraria from Biosense Webster Inc., Abbott Inc., Medtronic Inc., and Varian Inc. He has also received research grants from Biosense Webster Inc., and Abbott Inc. **Dr. Laurence D. Sterns** has received honoraria from Biosense Webster Inc., **Dr. Vijay S. Chauhan** has received consulting fees/honoraria and research support from Biosense Webster Inc. **Dr. Vidal Essebag** has received honoraria from Abbott, Biosense Webster, Boston Scientific, and Medtronic. The other authors have no disclosures related to this publication.

## Notes

### Competing Interest Statement

The authors have declared no competing interest.

### Clinical Trial

NCT02150902

### Author Declarations

Ethics Approval was obtained from the Ottawa Heart Research Institute and each separate hospital local ethics board.

